# Disease severity related to VOC dominance in unvaccinated SARS-CoV-2 positive adults without risk factors in Sweden

**DOI:** 10.1101/2021.12.23.21268040

**Authors:** Erik Wahlström, Daniel Bruce, Anna M Bennet Bark, Sten Walther, Håkan Hanberger, Kristoffer Strålin

## Abstract

Although the emerging SARS-CoV-2 variants of concern (VOC) have shown increasing transmissibility, their role for causing severe disease has not been fully clarified. Here, we studied changes in rates of hospitalisation and severe illness (subjection to high-flow nasal oxygen or admission to an intensive care unit during hospital stay) among all (n=685 891) unvaccinated SARS-CoV-2 positive adults without risk factors in Sweden from November 2020 to September 2021.

After adjustment for age, sex, and socio-economic factors, and with November 2020 (non-VOC period) as reference, the odds ratios (OR) for hospitalisation were 1.6-1.7 in March-May 2021 (Alpha VOC dominance) and 2.4-3.0 in June-September 2021 (Delta VOC dominance), and the ORs for severe illness were 1.8-2.1 in March-May 2021 and 3.1-4.7 in June-September 2021. This study shows that unvaccinated adults without risk factors, have had a gradually increased risk for hospital admission and severe illness when infected with the Alpha and Delta VOCs, respectively.

---

Since the autumn of 2020, certain linages of SARS-CoV-2 virus, i.e. variants of concern (VOC), have emerged and become dominating throughout the world. In Sweden, the Alpha VOC became dominating in the spring and the Delta COV became dominating during the summer of 2021 (Fig. 1A) [1]. In addition to altered transmissibility, the VOCs may harbour different virulence properties, e.g. the alpha VOC is associated with more severe disease than the first non-VOC lineages, as shown in previous studies [2, 3]. However, whether or not there is a further increased disease severity associated with the Delta VOC has not been fully clarified. Twohig et al. [4] found that the Delta VOC caused more hospitalisation and emergency care attendance than the Alpha VOC, although Taylor et al. [5] did not find any severity difference between the Delta period and the pre-Delta period among hospitalised COVID-19 patients.

**Fig. 1.**
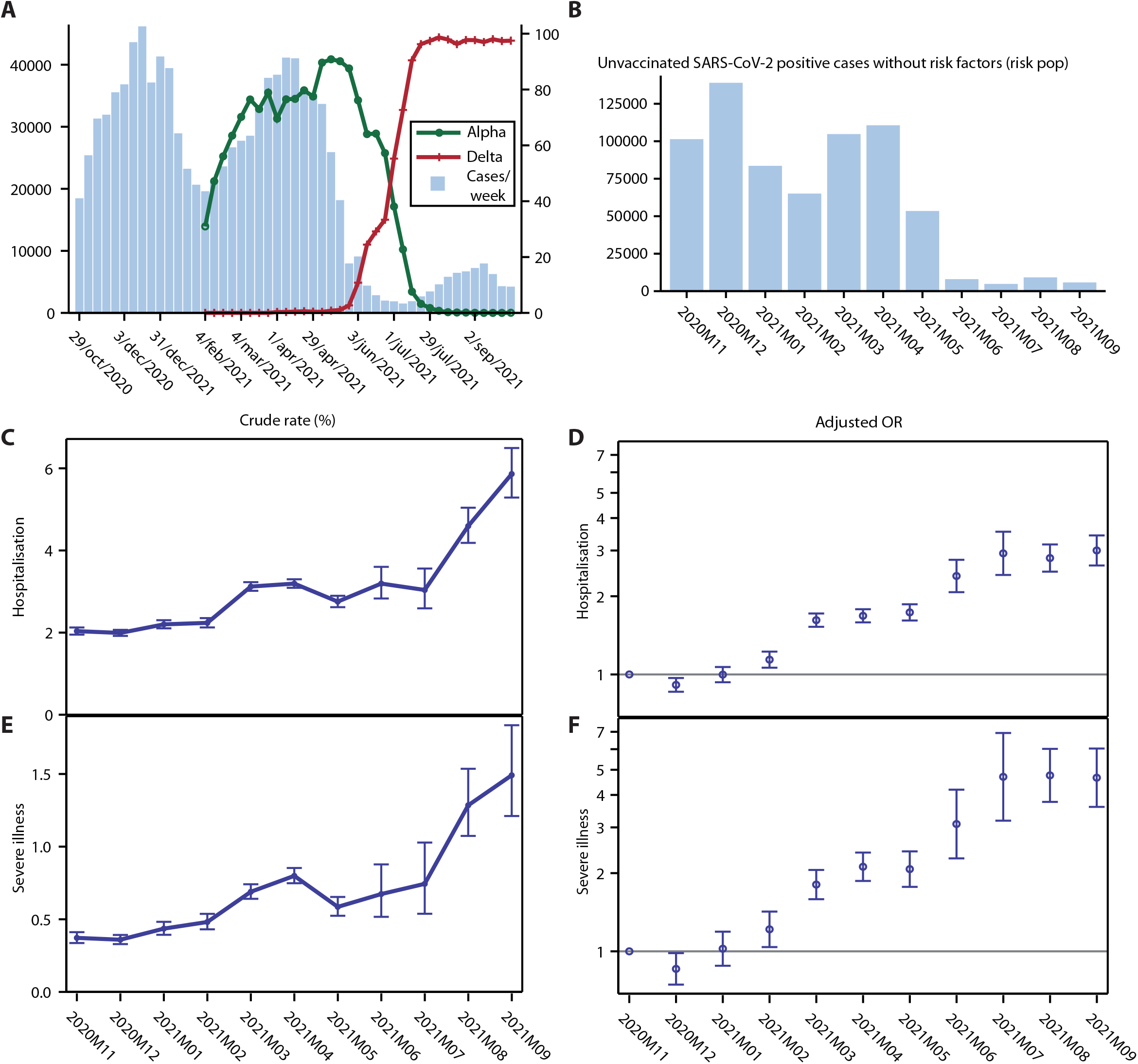
A, Distribution of SARS-CoV-2 positive individuals overall in Sweden between November 2020 and September 2021 (left Y-axis), with the proportion (%) of Alpha variant of concern (VOC) and Delta VOC indicated on right Y-axis [1]; B, Distribution of unvaccinated SARS-CoV-2 first-time positive individuals 20-69 years old, without comorbidity and without care dependency; C, Crude rate of hospitalisation among unvaccinated SARS-CoV-2 first-time positive individuals 20-69 years old, without comorbidity and without care dependency; D, Odds ratios (OR) for hospitalisation among unvaccinated SARS-CoV-2 first-time positive individuals 20-69 years old, without comorbidity and without care dependency, with adjustment for age, sex, and socio-economy, with November 2020 as reference; E, Crude rate of severe illness, defined as care with high-flow nasal oxygen or admission to an intensive care unit, among unvaccinated SARS-CoV-2 first-time positive individuals 20-69 years old, without comorbidity and without care dependency; F, OR for severe illness among unvaccinated SARS-CoV-2 first-time positive individuals 20-69 years old, without comorbidity and without care dependency, with adjustment for age, sex, and socio-economy, with November 2020 as reference.

In the present nationwide study, we aimed to study the rates of hospitalisation and severe illness among unvaccinated SARS-CoV-2 positive adults without risk factors over time and related to Alpha and Delta VOC dominance.

The study was conducted by the National Board of Health and Welfare in Sweden. It included all unvaccinated SARS-CoV-2 first-time test-positive individuals, 20-69 years old, without comorbidity and without care dependency and with positive test between November 2020 and September 2021. Nationwide register data on demographic factors, socio-economy (country of birth, education level, disposable income, main source of income), care dependency, comorbidity, hospitalisation, intensive care unit (ICU) admission, discharge codes, SARS-CoV-2 positivity data and anti-SARS-CoV-2 vaccination data were compiled, using the unique national personal identification number, as described previously [2, 6]. Individuals who were admitted to hospital, for any reason within 14 days after or 5 days before the first SARS-CoV-2 positive test, were considered hospitalised due to COVID-19. Those who either received high-flow nasal oxygen (according to a specific national discharge code) or were admitted to an ICU during their hospital stay were considered to have severe illness due to COVID-19.

Altogether, 685 891 unvaccinated SARS-CoV-2 positive adults without risk factors were included in the analysis. The distribution of these cases over the study period is shown in Fig. 1B. The characteristics of the study population changed over time. Patients admitted from November 2020 - February 2021 (second wave, non-VOC dominance, n=389 058), March-May 2021 (third wave, Alpha VOC dominance, n=268 954), and June-September 2021 (post-wave period, Delta VOC dominance, n=27 879) were female in 52%, 48%, and 48%, had median age 41 years, 41 years, and 32 years; were born outside of Sweden in 27%, 23%, and 41%; and belonged to the lowest income quintile in 14%, 13%, and 24%; respectively.

Overall, the study group had a hospitalisation rate of 2.6% and a severe illness rate of 0.55%. Fig. 1C and D show changes in hospitalisation rates over time. After adjustment for age, sex, and socio-economic factors (Fig 1D), three different levels of hospitalisation rates were noted. With November 2020 as reference the odds ratio (OR) for hospitalisation was 1.6 – 1.7 in March - May 2021 and 2.4 – 3.0 in June – September 2021. A similar pattern with three levels was noted regarding severe illness (Fig. 1F). With November 2020 as reference, the OR for severe illness was 1.8 – 2.1 in March - May 2021 and 3.1 – 4.7 in June – September 2021. The changes in ORs for hospitalisation and severe illness over time were consistent in all income quintiles (data not shown).

Our analysis, which shows increased hospitalisation and severe illness rates in individuals without risk factors, coinciding with changes in viral dominance, supports studies showing that Alpha VOC causes more severe disease than non-VOC [2, 3] and that Delta VOC causes more severe disease than Alpha VOC [4].

Some potentially important factors that we were unable to account for must be mentioned. First, we do not know if the correlation between true infected individuals and test-positive individuals was stable over the study period. However, throughout the study period the testing capacity in Sweden was high and the recommendation for testing was stable. Second, COVID-19 hospital strain was low in June-September 2021 which may have led to lower thresholds to admit individuals with mild COVID-19. However, the threshold to treat patients with high-flow nasal oxygen or admit patients to an ICU is most likely not correspondingly influenced by COVID-19 hospital pressure. Third, the changing characteristics of the unvaccinated population may have influenced the results. However, the study results were supported by the fact that the stepwise increases in hospitalisation and severe illness remained after adjustment for age, sex, and socio-economy in all income quintiles.

In the end of November 2021 the number of SARS-CoV-2 positive individuals started to increase in Sweden, with clear dominance of the Delta VOC [1]. The increased rates of hospitalisation and severe illness in unvaccinated SARS-CoV-2 positive individuals, shown in this study, must be considered in resource planning. It emphasizes the importance to decrease the proportion of unvaccinated individuals in the population.

The methodology used lends itself for continuous monitoring of hospitalisation and severity rates also after emergence of new VOC such as the Omicron VOC.

## Ethics

The study was approved by the Swedish Ethics Review Authority, Uppsala (Dnr 2020-04278).

## Data Availability

The data underlying this article cannot be shared publicly due to regulations under Swedish law. According to the Swedish Ethics Review Act, the General Data Protection Regulation, and the Public Access to Information and Secrecy Act, patient data can only be made available, after legal review, to researchers who meet the criteria for access to this type of confidential data. Requests regarding data in this report may be made to the corresponding author.

## Acknowledgments

This study was funded by Sweden’s National Board of Health and Welfare.

## References

1. Public Health Agency of Sweden. Statistik om SARS-CoV-2 virusvarianter av särskild betydelse. 2021 December 13, 2021 [cited December 13, 2021]; Available from: https://www.folkhalsomyndigheten.se/smittskydd-beredskap/utbrott/aktuella-utbrott/covid-19/statistik-och-analyser/sars-cov-2-virusvarianter-av-sarskild-betydelse/

2. Strålin K, Bruce D, Wahlström E, Walther S, Carnahan MRA, Andersson E, et al. Impact of the Alpha VOC on disease severity in SARS-CoV-2-positive adults in Sweden. J Infect 2021; S0163-4453(21)00448-5.

3. Cevik M, Mishra S. SARS-CoV-2 variants and considerations of inferring causality on disease severity. Lancet Infect Dis 2021; 21(11):1472–4.

4. Twohig KA, Nyberg T, Zaidi A, Thelwall S, Sinnathamby MA, Aliabadi S, et al. Hospital admission and emergency care attendance risk for SARS-CoV-2 delta (B.1.617.2) compared with alpha (B.1.1.7) variants of concern: a cohort study. The Lancet Infect Dis 2021; S1473-3099(21)00475-8.

5. Taylor CA, Patel K, Pham H, Whitaker M, Anglin O, Kambhampati AK, et al. Severity of Disease Among Adults Hospitalized with Laboratory-Confirmed COVID-19 Before and During the Period of SARS-CoV-2 B.1.617.2 (Delta) Predominance - COVID-NET, 14 States, January-August 2021.MMWR Morb Mortal Wkly Rep 2021 Oct 29;70(43):1513–1519.

6. Strålin K, Wahlström E, Walther S, Bennet-Bark AM, Heurgren M, Lindén T, et al. Mortality in hospitalized COVID-19 patients was associated with the COVID-19 admission rate during the first year of the pandemic in Sweden. Infect Dis (Lond) 2021; Oct 6:1–7.

